# Effects of Soybean-derived Phosphatidylserine on Working Memory and Motor Speed in Healthy Adults with Subjective Memory Complaints: A 12-Week Randomized, Placebo-Controlled Trial

**DOI:** 10.64898/2026.09.18.26363447

**Authors:** Yin Liu, Yunping Tang, Galex K.S. Neoh, Yangfan Lu, Tsuyoshi Takara, Su Jiang

## Abstract

Phosphatidylserine (PS) is a functional phospholipid widely studied for cognitive support, yet evidence in healthy adults with subjective cognitive decline remains limited. This 12-week randomized, double-blind, placebo-controlled trial evaluated the safety and cognitive effects of 300 mg/day of soybean-derived PS in 70 adults aged 35–65 years reporting subjective memory complaints. PS was well tolerated, with no adverse events or clinically significant changes in safety parameters. In the prespecified primary analysis, the primary endpoint of composite memory score did not differ significantly between groups at week 12. Exploratory secondary analyses suggested possible domain-specific effects, including favorable changes in working memory (*p* = 0.037) and motor speed (*p* = 0.005), with the latter appearing to vary according to blood fatty-acid profiles. These findings should be interpreted cautiously and require confirmation in future studies designed to evaluate these domains and potential nutritional moderators.

## 1. Introduction

Cognitive health has become an increasingly important public concern, driven by demographic aging, and stressor exposures in everyday living including rising occupational stress, chronic sleep insufficiency, and heightened psychological demands [1]. Although Alzheimer’s disease (AD) and mild cognitive impairment (MCI) remain the most widely recognized neurological disorders, many individuals experience subjective cognitive decline (SCD) – a self-perceived deterioration in memory or cognitive efficiency despite normal performance on standardized tests [2]. SCD is not a clinical diagnosis but is considered an early symptomatic stage that may precede MCI or AD [3]. SCD is increasingly recognized as a clinically relevant at-risk state rather than merely a benign subjective experience. A recent systematic review and meta-analysis of longitudinal studies reported that the mean conversion rate from SCD to any cognitive deterioration was 19.8%, including 7.3% to all-cause dementia and 4.9% to Alzheimer’s disease, although progression risk is heterogeneous across individuals. These findings support the view that SCD may represent an early symptomatic stage of later objective cognitive decline in at least a subset of affected individuals [4]. The public health burden of SCD is substantial. In a nationwide U.S survey, one in nine adults aged ≥ 45 years reported SCD, yet fewer than half had discussed their concerns with a healthcare professional [5]. Globally, SCD prevalence ranges from 17 – 46%, with higher rates among older adults and individuals experiencing psychological stress or mood symptoms [6]. Beyond concerns about memory, SCD is associated with functional limitations, reduced social participation, and elevated psychological distress, including fear of dementia [7,8]. Because many individuals with SCD typically do not seek medical evaluation, a large proportion of those experiencing cognitive concerns may instead turn to over-the-counter nutritional supplements marketed for cognitive health support.

Phosphatidylserine (PS) is one of the most widely used supplement ingredients for cognitive support. It is an essential phospholipid abundant in neuronal membranes, where it contributes to membrane fluidity, neurotransmitter release, and synaptic signaling [9,10]. In the adult human brain, PS constitutes approximately 12–14% of cortical phospholipids and up to 20% in white matter. Aging is associated with a decline in endogenous PS levels, accompanied by reduced membrane integrity and synaptic function [11,12]. Supplementation with exogenous PS has been shown to be bioavailable, capable of crossing the blood-brain barrier, and able to incorporate into neuronal membranes, thereby potentially mitigating age-related declines in neural structure and function [10]. Mechanistically, PS plays multiple roles in neuronal physiology. As the primary anionic phospholipid of the inner membrane leaflet, PS provides essential docking sites for signaling proteins involved in neuronal survival and synaptic plasticity, including Akt, PKC, and Raf-1 [13,14]. PS also regulates Ca^2+^-dependent synaptic vesicle fusion [15] and modulates glutamatergic receptor function, partially restoring age-related deficits in NMDA and AMPA receptor signaling [16]. These structural and functional properties have generated sustained interest in PS as a nutritional compound for cognitive enhancement.

Clinical research on PS has demonstrated cognitive benefits in several populations, particularly older adults with age-related memory impairment or MCI. Studies report improvements in memory, learning, executive functioning, and mental flexibility following PS supplement [17–19]. More recently, a randomized double-blind, placebo-controlled trial in Chinese older adults with MCI reported cognitive benefits from a multi-ingredient supplement containing phosphatidylserine, α-linolenic acid, ginkgo flavonoids, and B vitamins over 12 months. These findings further support interest in PS-containing interventions for cognitive health, while also highlighting the need to better understand the effects of soybean-derived PS as a defined ingredient in healthier, younger populations with subjective cognitive concerns [19]. Despite this promising evidence, most trials have a limited focus on the older adult population. Considerably fewer studies have examined PS supplementation in healthy middle-aged adults experiencing subjective cognitive concerns. The cognitive and physiological profiles of these individuals differ from those with MCI or AD, and results from older clinical population cannot be directly extrapolated to younger adults in the preclinical phase of cognitive decline [2]. This gap leaves a substantial portion of the supplement-using population underrepresented in controlled clinical research. Commercially available PS supplements have also transitioned over time. Early formulations used bovine-derived PS, but concerns surrounding bovine spongiform encephalopathy led to regulatory restrictions and a subsequent shift towards plant-derived sources, particularly soybean-derived PS [20]. Soybean-derived PS is now the predominant form used in cognitive health products and has been shown to exert cognitive benefits similar to animal-derived PS. Yet, there remains limited randomized controlled trial data evaluating the cognitive benefits specifically in healthy adults with cognitive complaints and SCD.

The present randomized, double-blind, placebo-controlled study was designed to address this gap by evaluating the safety and effects of daily supplementation with soybean-derived PS in healthy adults aged 35–65 years reporting subjective cognitive concerns. Cognitive function was assessed using Cognitrax^®^ computerized testing system, which is a validated assessment providing a multidomain evaluation of memory, attention, executive function, processing speed, and motor speed. In addition to conventional analyses, the study applied multilevel statistical models that can account for repeated observation over time and the potential moderating influence of baseline cognitive status and blood fatty-acid ratio.

## 2. Material and methods

### 2.1. Study design and ethical approval

This study was a randomized, double-blind, placebo-controlled, parallel-group trial conducted over 12 weeks to evaluate the safety and exploratory cognitive effects of PS supplementation in healthy adults experiencing subjective memory decline. The trial was approved by the Institutional Review Board of Medical Corporation Seishinkai, Takara Clinic (Tokyo, Japan) on June 15, 2023 (No. 2306-06788-0019-10-TC). The protocol adhered to the principles of the Declaration of Helsinki (2013) and Japan’s Ethical Guidelines for Medical and Health Research Involving Human Subjects. All participants provided written informed consent prior to enrollment. This trial was prospectively registered at ClinicalTrial.gov (NCT05962008) and the UMIN Clinical Trials Registry (UMIN000051592).

The original registered trial included a third intervention arm receiving fish roe-derived PS. However, the present manuscript was designed to address a source-specific question, namely the safety and cognitive effects of soybean-derived PS in healthy adults aged 35–65 years with subjective cognitive concerns. Fish roe-derived PS differs materially from soybean-derived PS in source composition and molecular characteristics, including its fatty acid profile. Therefore, including both PS preparations in the present manuscript would change the scientific question from a placebo-controlled evaluation of a defined soybean-derived PS intervention to a comparative source analysis across distinct PS preparations. Because the objective of this report was to provide a focused evaluation of soybean-derived PS, the predominant commercially used PS form in global cognitive health products, the fish roe-derived PS arm was excluded from the present efficacy analysis. This restriction was made to preserve interpretive clarity around one defined intervention rather than to selectively emphasize favorable findings. The existence of the third arm is disclosed here for transparency. This restriction of the analysis set does not affect the original ethical approval, trial registration, safety monitoring, or trial conduct.

### 2.2. Sample size and participants

Because no previous clinical study had evaluated the composite memory score of Cognitrax^®^ at 12 weeks after consumption of the test food, the sample size could not be based on a directly comparable prior study using the same endpoint. Therefore, the sample size calculation assumed a large effect size (Cohen’s d = 0.80), based on Cohen’s suggestion, with a two-sided significance level of 0.05 and statistical power of 0.90. On this basis, the required number of participants to be analyzed was 105 (35 per group in the original three-arm design). To account for anticipated dropout and protocol noncompliance during the trial period, the planned enrollment was increased to 114 participants (38 per group). Participants were recruited through online study announcements. Inclusion criteria were: (1) healthy men and women aged 35–65 years; (2) who reported subjective memory concerns; (3) a Mini-Mental State Examination (MMSE) score of ≥ 24 at screening, used as a protocol-defined threshold to exclude participants with clear overall cognitive impairment; and (4) completion of the Cognitrax^®^ cognitive test battery during screening. Key exclusion criteria comprised: (1) regular intake of supplements or foods known to enhance cognitive function (e.g., docosahexaenoic acid (DHA), eicosapentaenoic acid (EPA), ginkgo leaf extract); (2) use of prescription medications or dietary supplements; (3) a history of mental health issues (e.g., depression, ADHD); (4) treatment for chronic diseases such as liver disease, kidney disease, or hypertension; (5) pregnancy or breastfeeding; and (6) known allergies to the test food products. A total of 210 individuals were screened, and 114 were enrolled following eligibility assessment. All participants provided informed consent prior to participation.

### 2.3. Randomization and blinding

The original trial randomized eligible participants into a 1:1:1 ratio to the soybean-derived PS (BioPS^®^), fish roe-derived PS (DHAPS^®^) or placebo groups. An independent allocation controller performed the randomization using a computer-generated sequence. Allocation was stratified by age (≥ 50 years vs. < 50 years) and the baseline (EPA + DHA)/ rachidonic acid (AA) ratio (≥ median vs. < median) to ensure group balance. The study was double-blinded; participants, investigators, and all study staff remained unaware of the group assignments until the trial was concluded, and the database was locked (e.g., all data were verified and finalized). This procedure ensures that no further changes can be made to the dataset before unblinding. Unblinding and statistical analyses were conducted only after the database lock to ensure the integrity and objectivity of the results.

### 2.4. Intervention

Participants were instructed to consume four capsules daily after breakfast with warm water for 12 weeks. In the original three-arm design, participants received either 300 mg/ day of BioPS^®^, 300 mg/ day of DHAPS^®^, or a placebo consisting of microcrystalline cellulose. All capsules were identical in appearance, shape, color, smell, and taste to maintain blinding. The detailed formulation of BioPS^®^ and placebo is provided in Table 1. The composition of the DHAPS^®^ formulation is not reported here because it falls outside the source-specific scope of the present analysis. Consistent with the study objectives described in Section 2.1, the present manuscript reports only the BioPS^®^ and placebo groups.

**Table 1.** Composition of the BioPS^®^ and placebo capsules administered during the 12-week intervention.

| Intervention | Form | Microcrystal<br>line<br>cellulose<br>(mg/capsule) | 60%<br>BioPS®<br>powder<br>(mg/capsule) | Rosemary<br>extract<br>(mg/capsule) | Sodium<br>erythorbate<br>(mg/capsule) | Magnesium<br>stearate<br>(mg/capsule) |
| --- | --- | --- | --- | --- | --- | --- |
| Placebo | Capsule | 293.25 mg | – | 2.10 mg | 0.15 mg | 4.50 mg |
| BioPS® | Capsule | 168.25 mg | 125.00 mg | 2.10 mg | 0.15 mg | 4.50 mg |

Both formulations included identical excipients to ensure matched capsule appearance and handling.

The 12-week intervention duration was selected in accordance with the original study protocol and was considered sufficient for an initial evaluation of safety and short-term cognitive effects, while remaining consistent with several previous PS clinical trials using 12-week supplementation periods [11,17].

### 2.5. Safety and efficacy assessment of PS intervention

Safety and tolerability were evaluated as primary outcomes of the study. The primary safety endpoint was the incidence of all adverse events. Events classified as adverse included any newly occurring symptoms, worsening of existing conditions, or clinically relevant abnormalities identified during safety evaluations. Secondary safety endpoints included the proportion of participants whose urinalysis or peripheral blood examination results shifted from within the normal reference range at baseline to outside the range at week 12. Other safety measures included anthropometric data (body weight, BMI), vital signs (systolic and diastolic blood pressure), standard urinalysis (protein, glucose, pH, occult blood), and comprehensive peripheral blood panels. The blood panels assessed hematology (WBC, RBC, hemoglobin, hematocrit, platelets) and biochemistry, including markers for liver function (AST, ALT, γ-GTP, total bilirubin), kidney function (urea nitrogen, creatinine, uric acid), and metabolic status (total protein, electrolytes, serum amylase, lipid panel, glucose, HbA1c). Blood fatty-acid fractions, including AA, EPA, dihomo-γ-linolenic acid (DHLA), DHA, and the derived ratios EPA/AA, DHA/AA, and (EPA + DHA)/AA, were also measured in fasting blood samples obtained at baseline and week 12.

Cognitive function was assessed using the Cognitrax^®^ computerized testing system. The primary efficacy endpoint was the standardized score of composite memory at 12 weeks, representing a composite index derived from standardized scores of verbal and visual memory. Secondary efficacy endpoints included the change from baseline in composite memory and the 12-week scores and change from screening (baseline) for 14 other cognitive domains, including working memory and motor speed. Working memory was defined as the ability to retain information while simultaneously manipulating and integrating additional information to achieve a cognitive objective (e.g., performing mental calculations or processing complex sentences). Motor speed was assessed using the Finger Tapping Test, which measures fine psychomotor function by evaluating the speed at which participants repeatedly tap a designated key.

### 2.6. Statistical analysis

All analyses were performed comparing the BioPS^®^ group to the placebo group. Incidence rates for adverse events and laboratory shifts were compared using Fisher’s exact test or the chi-squared test, as appropriate.

The prespecified primary efficacy analysis was conducted using analysis of covariance (ANCOVA), consistent with the original study protocol. ANCOVA models compared week-12 cognitive endpoints between groups while adjusting for the corresponding baseline cognitive score. The primary efficacy endpoint was the week-12 standardized composite memory score, adjusted for baseline composite memory score. Secondary cognitive endpoints were analyzed in the same manner.

In addition to the prespecified ANCOVA, exploratory secondary analyses were conducted using mixed-effects models to examine whether treatment-related differences could be detected in change patterns over time for selected cognitive domains of interest. These exploratory analyses were limited to working memory and motor speed, given their relevance to subjective cognitive concerns and the possibility that subtle, domain-specific responses may be present in a healthy, high-functioning population. The placebo group was used as the reference category.

The mixed-effects models included fixed effects for group, time, and the group × time interaction. A time-varying covariate, the blood (EPA + DHA)/AA ratio, was included in exploratory models evaluating potential moderation by nutritional status. Additional baseline covariates included age, sex, education, BMI, and baseline composite memory score. Where higher-order interaction terms were not statistically significant, they were removed to obtain a more parsimonious model.

All mixed-effects analyses were conducted using SAS PROC MIXED with restricted maximum likelihood estimation. These exploratory analyses were intended to generate hypotheses regarding response heterogeneity and potential nutritional moderation and were not considered a replacement for the prespecified primary ANCOVA. A two-tailed p-value < 0.05 was considered statistically significant.

## 3. Results

### 3.1. Study population

A total of 216 individuals were screened for eligibility, of whom 114 were enrolled and randomized into the original three-arm trial (Fig. 1). For the present analysis, 70 participants who completed the trial were included, comprising the BioPS^®^ group (*n* = 33) and the placebo group (*n* = 37). Six participants were excluded from the analysis (five participants from the BioPS^®^ group, one participant from the placebo group) after being lost to follow-up, as they did not return for the examination at 12 weeks. No withdrawals were related to safety concerns.

**Fig. 1.**
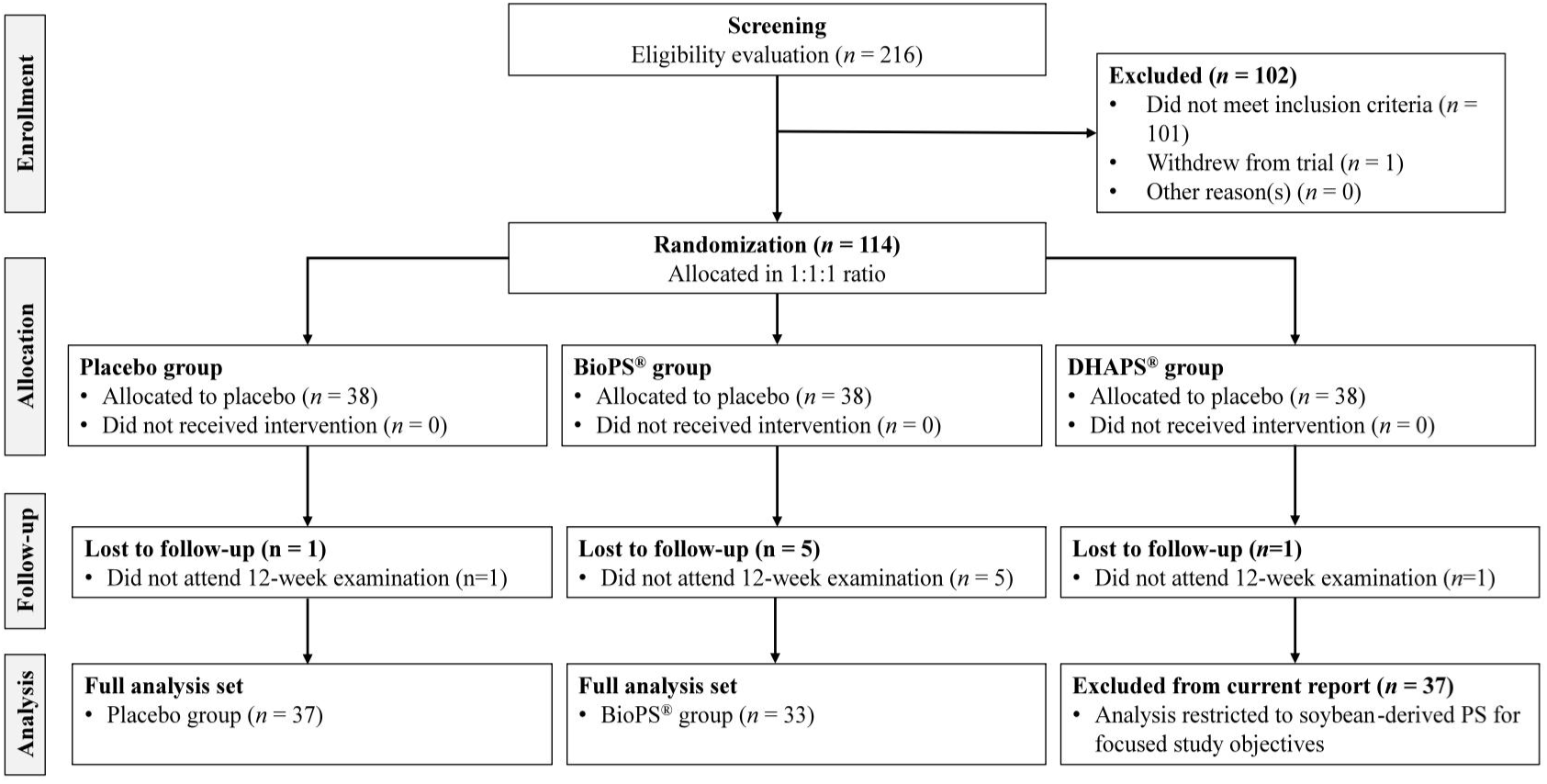
Flowchart of participant recruitment, randomization, follow-up and analysis for the 12-week intervention.

Baseline demographic and clinical characteristics were generally comparable between BioPS^®^ and placebo groups. Mean age, sex distribution, BMI, education level and baseline composite memory score did not differ statistically between groups (Table 2 and Table 3). However, a statistically significant difference was observed in baseline motor speed (*p* = 0.03), which was accounted for by inclusion of baseline motor speed as a covariate in subsequent efficacy models. The rate of compliance in consuming the prescribed intervention product was high for both groups, with mean compliance of 99.9 ± 1.0% in the placebo group and 99.5 ± 2.1% in the BioPS^®^ group. Overall, the randomization and selection process resulted in two groups that were well balanced at baseline for comparative safety and efficacy evaluation of BioPS^®^.

**Table 2.**
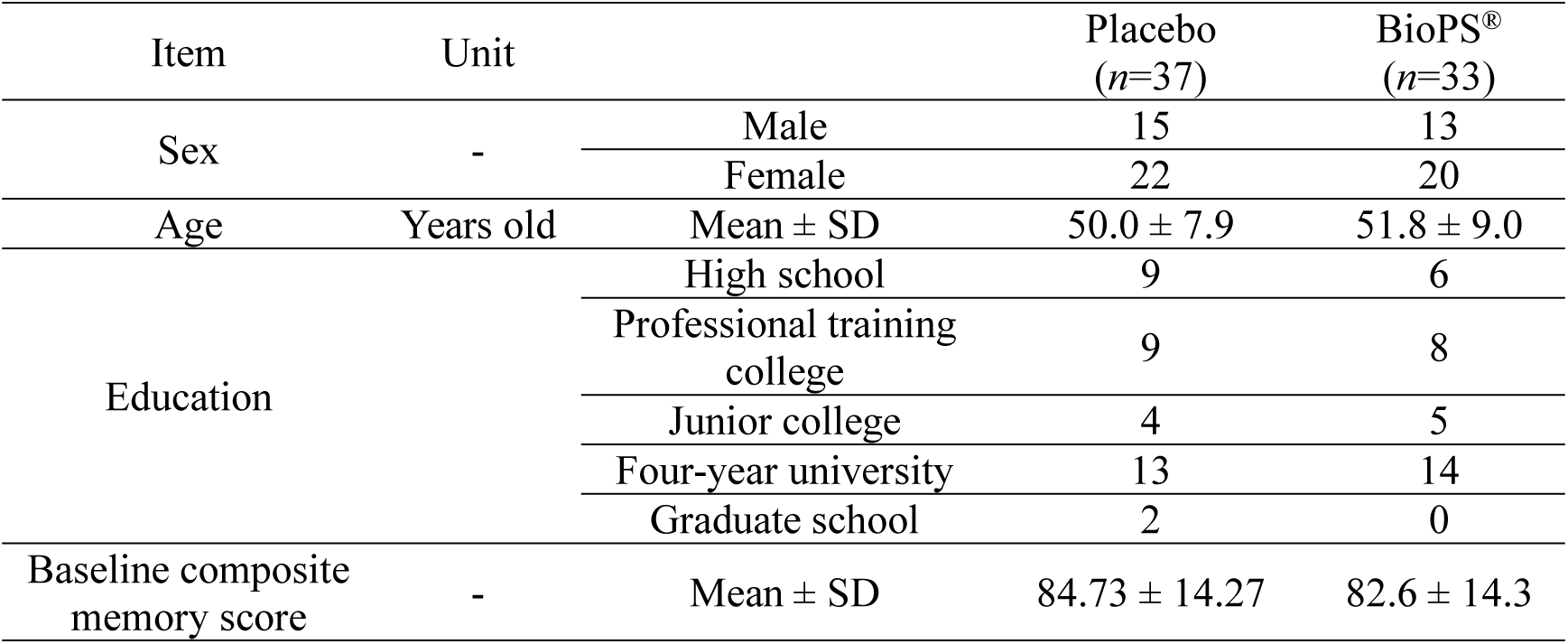
Baseline demographic and cognitive characteristics of participants in the BioPS® and placebo groups.

| Item | Unit |  | Placebo<br>(n=37) | BioPS®<br>(n=33) |
| --- | --- | --- | --- | --- |
| Sex | - | Male | 15 | 13 |
|  |  | Female | 22 | 20 |
| Age | Years old | Mean ± SD | 50.0 ± 7.9 | 51.8 ± 9.0 |
| Education |  | High school | 9 | 6 |
|  |  | Professional training college | 9 | 8 |
|  |  | Junior college | 4 | 5 |
|  |  | Four-year university | 13 | 14 |
|  |  | Graduate school | 2 | 0 |
| Baseline composite memory score | - | Mean ± SD | 84.73 ± 14.27 | 82.6 ± 14.3 |

**Table 3.** Anthropometric measurements and vital signs at baseline and week 12 for the BioPS^®^ and placebo groups.

| Parameter | Time point | Placebo (*n* = 37)<br>Mean ± SD | BioPS® (*n* = 33)<br>Mean ± SD |
| --- | --- | --- | --- |
| Body weight (kg) | Baseline | 61.4 ± 12.3 | 62.6 ± 14.9 |
|  | Week 12 | 62.4 ± 12.2 | 63.2 ± 15.6 |
| Body height (cm) | Baseline | 164.6 ± 9.6 | 163.2 ± 9.6 |
|  | Week 12 | 164.0 ± 9.7 | 162.9 ± 9.6 |
| BMI (kg/m²) | Baseline | 22.5 ± 3.1 | 23.3 ± 3.6 |
|  | Week 12 | 22.9 ± 3.1 | 23.6 ± 3.8 |
| Systolic blood pressure (mmHg) | Baseline | 122.0 ± 15.5 | 121.1 ± 16.6 |
|  | Week 12 | 125.6 ± 16.8 | 123.8 ± 16.3 |
| Diastolic blood pressure (mmHg) | Baseline | 78.8 ± 13.2 | 75.1 ± 11.0 |
|  | Week 12 | 81.7 ± 12.3 | 79.8 ± 13.1 |

All between-group differences were non-significant (*p* > 0.05).

### 3.2. Safety evaluation of PS

No adverse events were reported in either the BioPS^®^ group or the placebo group throughout the 12-week intervention period. Anthropometric values and vital signs, including body weight, BMI, and blood pressure remained stable from baseline to week-12 in both groups (Table 3). Mean changes over time were not statistically significant within groups or between groups.

Urinalysis results, including protein, glucose, pH and occult blood, showed no clinically meaningful changes after 12 weeks of intervention (Table 4). The proportion of participants whose values shifted from within the reference range at baseline to outside the range at week 12 was low and comparable between BioPS^®^ and placebo groups. No pattern suggestive of urinary abnormalities was observed.

**Table 4.**
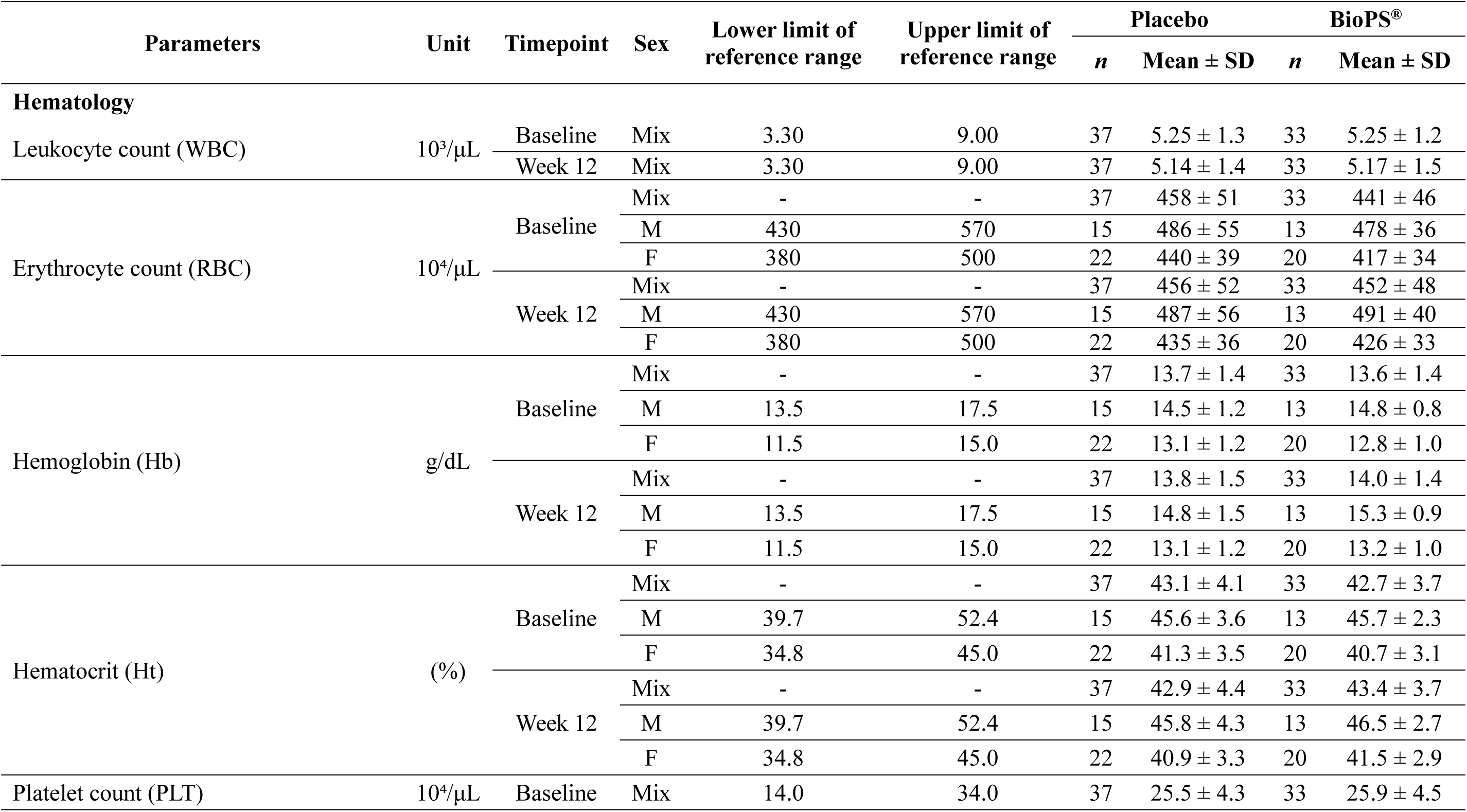
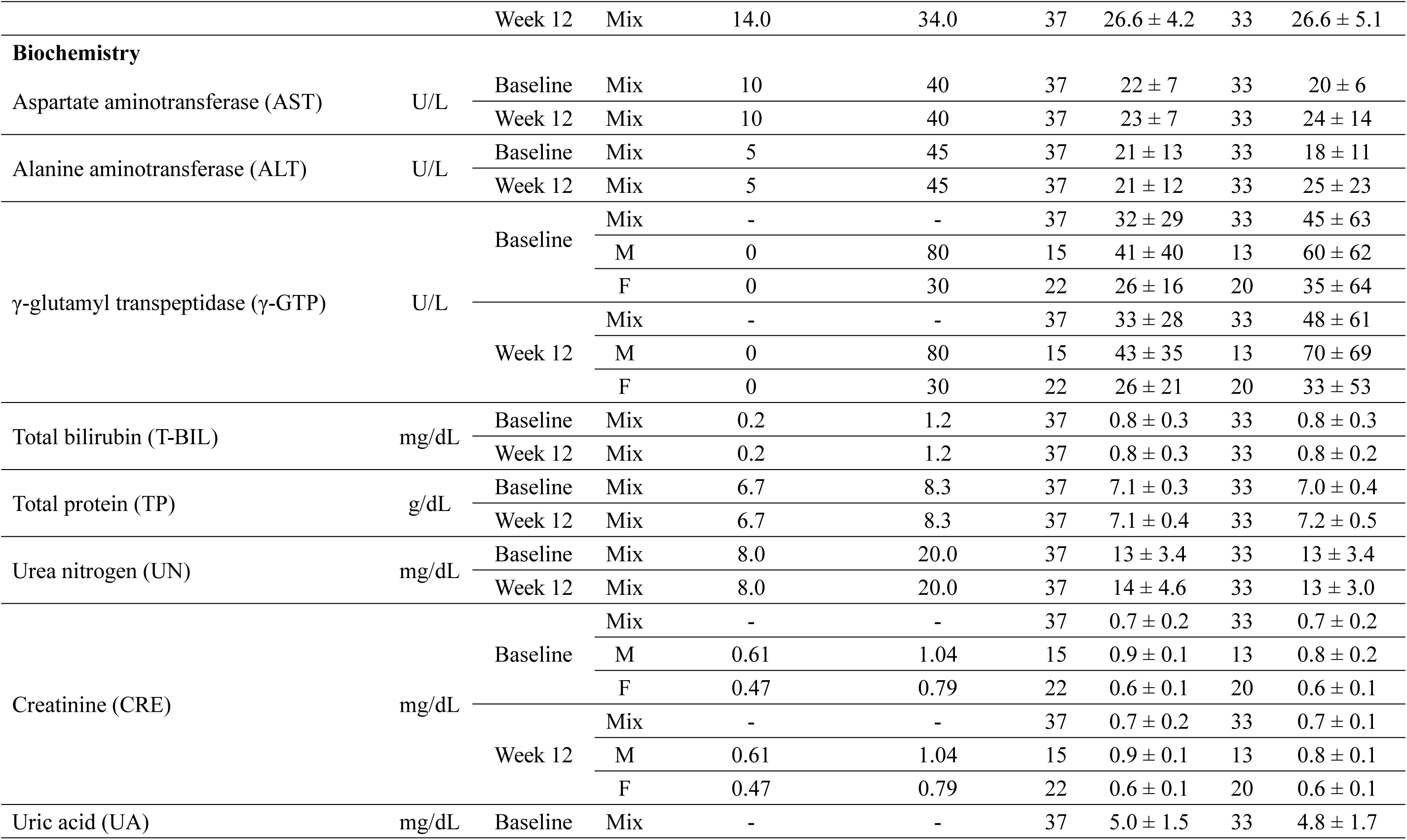
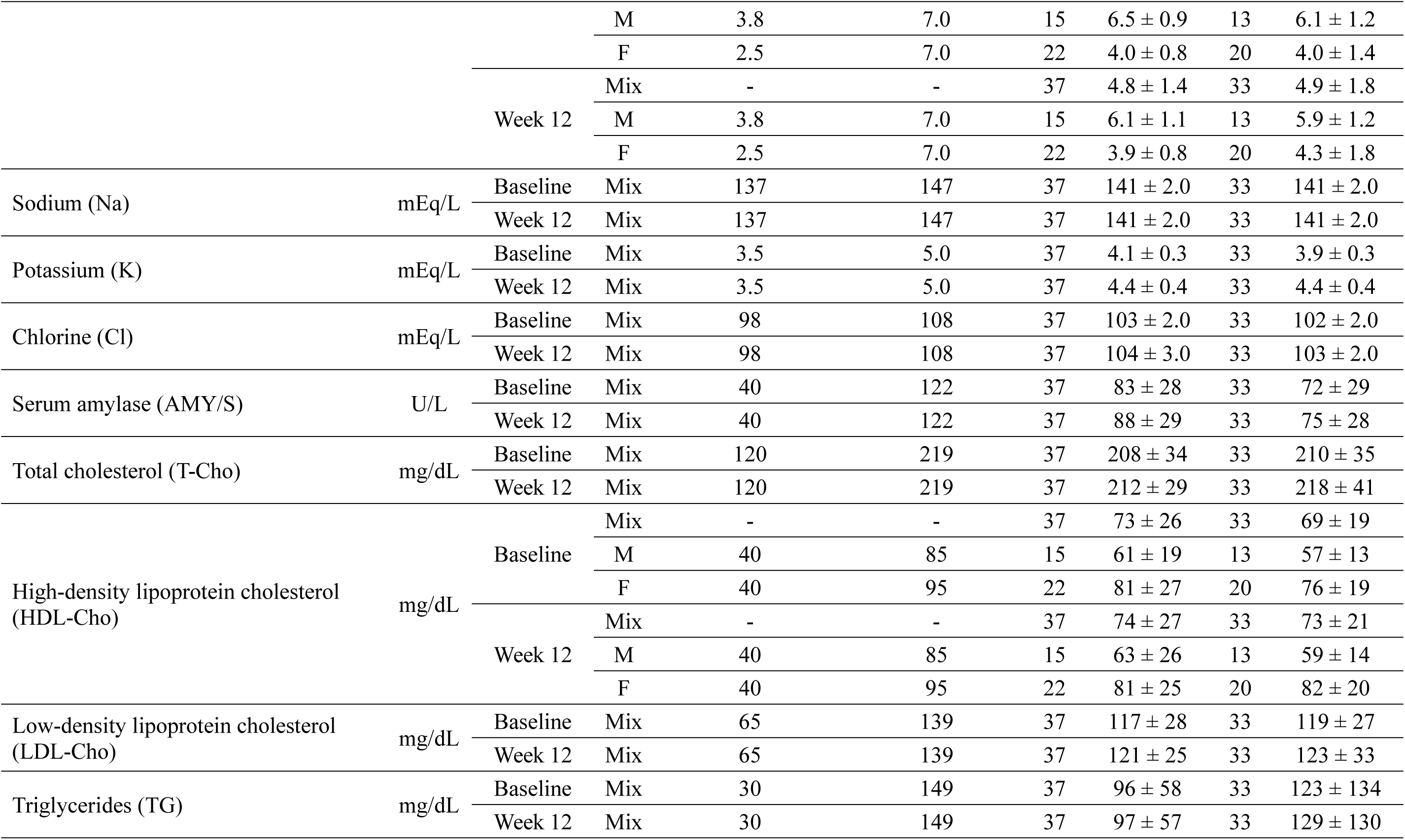
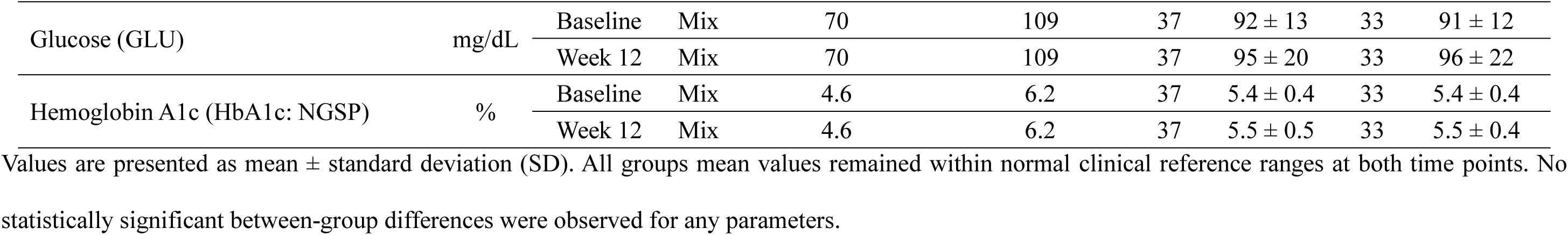
Hematological and biochemical parameters at baseline and week 12 in the BioPS^®^ and placebo groups.

A comprehensive peripheral blood examination found that all mean hematological and biochemical parameters remained stable and within normal clinical reference ranges in both groups from baseline to week 12 (Table 4). These included markers for liver function (AST, ALT, bilirubin), kidney function (creatinine, urea nitrogen), and metabolic health (lipids, glucose). Although a few participants exhibited isolated shifts from in-range to out-of-range values, these occurrences were sporadic, showed no discernible pattern, and did not differ significantly between groups. Importantly, none of the observed laboratory shifts were clinically significant upon review.

Taken together, these safety assessments support the safety of daily BioPS^®^ consumption at administered dosage of 300 mg/day over the 12-week intervention period.

### 3.3. BioPS^®^ effects on cognitive function

Using the prespecified ANCOVA approach, no statistically significant between-group differences were observed for the primary efficacy endpoint, composite memory score, at week 12 after adjustment for baseline values. Likewise, no significant between-group differences were detected for working memory or motor speed in the primary ANCOVA framework. Mean changes in other cognitive domains were generally small and comparable between the BioPS^®^ and placebo groups.

Because subjective cognitive concerns in otherwise healthy adults may manifest as subtle and domain-specific changes rather than broad cognitive shifts, exploratory secondary analyses were conducted for selected cognitive domains of interest. These analyses were intended to examine whether treatment-related differences in change over time, and potential moderation by nutritional status, could be detected beyond the prespecified endpoint comparison.

Exploratory subgroup analyses suggested that treatment effects may have been more apparent among participants with lower baseline cognitive or nutritional status. For example, among participants with baseline composite memory scores below the median, the BioPS^®^ group showed greater improvement in sustained attention than the placebo group. Similarly, participants with lower baseline fatty-acid-related status showed larger improvements in selected cognitive outcomes. These findings should be interpreted cautiously, as these subgroup analyses were exploratory.

Exploratory mixed-effects models were then applied to working memory and motor speed to examine group differences in change over time and the possible moderating role of the blood (EPA + DHA)/AA ratio (Table 5). For working memory, the reduced model including the time × group interaction showed a significant interaction term (β = 7.594, SE = 3.588, p = 0.037), suggesting a more favorable change pattern over time in the BioPS^®^ group relative to placebo. For motor speed, the model including the time × group × fatty-acid ratio interaction showed a significant three-way interaction (β = -55.635, SE = 19.223, p = 0.005), suggesting that the effect of BioPS^®^ on change in motor speed varied according to changes in the blood (EPA + DHA)/AA ratio over the intervention period.

**Table 5.** Multilevel growth curve model estimates for cognitive outcomes of working memory and motor speed trajectories over the 12-week intervention.

| Fixed Effect | Working memory |  |  |  | Motor speed |  |  |  |  |  |
| --- | --- | --- | --- | --- | --- | --- | --- | --- | --- | --- |
|  | Model 1 |  | Model 2 |  | Model 1 |  | Model 2 |  | Model 3 |  |
|  | Est ± SE | *p*-value | Est ± SE | *p*-value | Est ± SE | *p*-value | Est ± SE | *p*-value | Est ± SE | *p*-value |
| Intercept | 85.449 ± 13.059 | <0.0001 | 87.765 ± 13.124 | <0.0001 | 69.573 ± 20.258 | 0.001 | 71.027 ± 20.340 | 0.001 | 73.777 ± 20.818 | 0.001 |
| Time | 0.741 ± 1.472 | 0.616 | -2.926 ± 2.484 | 0.242 | -2.095 ± 2.494 | 0.403 | -5.685 ± 4.237 | 0.183 | -17.116 ± 7.560 | 0.026 |
| BioPS® | 0.643 ± 2.593 | 0.805 | -2.919 ± 3.094 | 0.348 | -0.165 ± 4.020 | 0.967 | -0.503 ± 4.943 | 0.919 | -4.358 ± 11.554 | 0.707 |
| Time × BioPS® | — | — | 7.594 ± 3.588 | 0.037 | — | — | 0.451 ± 6.112 | 0.941 | 33.465 ± 14.076 | 0.019 |
| Time × Acid ratio | — | — | — | — | — | — | — | — | 23.961 ± 10.251 | 0.021 |
| Acid ratio × BioPS® | — | — | — | — | — | — | — | — | 8.992 ± 15.532 | 0.564 |
| Time × Acid ratio × | — | — | — | — | — | — | — | — | -55.635 ± 19.223 | 0.005 |
| Covariates |  |  |  |  |  |  |  |  |  |  |
| Acid ratio | 3.342 ± 3.555 | 0.349 | 3.371 ± 3.552 | 0.345 | -3.389 ± 5.650 | 0.550 | -3.986 ± 5.646 | 0.482 | -11.079 ± 8.666 | 0.204 |
| Age | -0.221 ± 0.126 | 0.083 | -0.229 ± 0.126 | 0.072 | -0.197 ± 0.196 | 0.316 | -0.194 ± 0.195 | 0.324 | -0.225 ± 0.200 | 0.263 |
| Sex | -2.038 ± 2.450 | 0.407 | -1.919 ± 2.453 | 0.436 | 9.862 ± 3.796 | 0.011 | 9.801 ± 3.794 | 0.011 | 9.796 ± 3.870 | 0.013 |
| Education | 1.706 ± 0.893 | 0.059 | 1.678 ± 0.894 | 0.064 | -0.795 ± 1.386 | 0.568 | -0.806 ± 1.385 | 0.562 | -0.990 ± 1.419 | 0.487 |
| BMI | 0.258 ± 0.366 | 0.481 | 0.249 ± 0.366 | 0.498 | -0.115 ± 0.568 | 0.840 | -0.095 ± 0.567 | 0.867 | -0.088 ± 0.577 | 0.879 |
| Composite memory | 0.215 ± 0.079 | 0.008 | 0.216 ± 0.079 | 0.007 | 0.528 ± 0.122 | <0.0001 | 0.530 ± 0.122 | <.0001 | 0.549 ± 0.125 | < .0001 |
| Random Effect |  |  |  |  |  |  |  |  |  |  |
| Intercept variance | 63.408 ± 18.949 | 0.0004 | 65.186 ± 18.960 | 0.0003 | 123.290 ± 46.699 | 0.004 | 125.160 ± 46.534 | 0.004 | 149.670 ± 47.591 | 0.001 |
| Residual | 118.030 ± 16.124 | <0.0001 | 115.110 ± 15.885 | <0.0001 | 339.710 ± 46.063 | <0.0001 | 335.200 ± 45.835 | <0.0001 | 306.470 ± 42.425 | < 0.0001 |

| **Model Fit** |  |  |  |  |  |
| --- | --- | --- | --- | --- | --- |
| -2 Log-likelihood | 1737.5 | 1724.6 | 1941.6 | 1927.4 | 1904 |
| AIC, BIC | 1741.5, 1747.0 | 1728.6, 1734.1 | 1945.6, 1951.1 | 1931.4, 1936.9 | 1908.0, 1913.5 |

Time was coded as 0 = baseline and 1 = week 12. The (EPA + DHA)/AA fatty-acid ratio was included as a time-varying covariate. For working memory, the three-way interaction (time × group × acid ratio) was not significant and was removed for model parsimony. Significant effects ( $p < 0.05$ ) are indicated in bold. Est = parameter estimate; SE = standard error; AIC = Akaike Information Criterion; BIC = Bayesian Information Criterion.

These mixed-effects findings should be interpreted as exploratory and hypothesis-generating, particularly because the prespecified primary ANCOVA did not detect significant between-group differences for these outcomes.

## 4. Discussion

The present 12-week, randomized, double-blind, placebo-controlled clinical trial evaluated the safety and efficacy of 300 mg/day of BioPS^®^ in healthy, middle-aged adults with subjective memory complaints. The primary safety endpoint was achieved. BioPS^®^ was well-tolerated, with no adverse events reported in either treatment groups, and no clinically significant changes were observed in anthropometric measurements, urinalysis parameters, or comprehensive hematological and biochemical laboratory panels. These findings are consistent with previous randomized controlled trials reporting that daily supplementation with soybean-derived PS is safe and well tolerated across a range of doses and study durations [17,18,21].

The findings exploring efficacy of BioPS^®^ were more nuanced. The prespecified primary efficacy endpoint, composite memory score, did not differ significantly between groups in the initial ANCOVA. However, secondary exploratory subgroup analyses suggested possible differences favoring BioPS^®^ in working memory (*p* = 0.077) and sustained attention (*p* = 0.074). These findings suggested the possibility of domain-specific effects in working memory and sustained attention. To further examine these observations, exploratory mixed-effects analyses were conducted to assess change patterns over time. This exploratory mixed-effects analysis suggested that BioPS^®^ was associated with favorable changes in working memory (*p* = 0.037) and motor speed (*p* = 0.005) compared with placebo (Table 5). This pattern of findings, where the primary analyses conducted in a broad, heterogeneous population do not reach significance, but additional analyses within a more homogeneous subgroups or using longitudinal modeling suggest possible domain-specific effects is consistent with observation from previous PS trials. For example, Kato-Kataoka et al. [18] reported that PS-related improvements were most evident in participants with lower baseline memory scores, and Jorissen et al. [21] found benefits primarily among individuals with elevated subjective cognitive complaints rather than in the entire study population. These collectively suggest that the cognitive benefits of PS are more apparent in specific subpopulations, particularly those with sub-optimal baseline status. Because these observations arose from exploratory analyses rather than the prespecified primary efficacy analysis, they should be interpreted cautiously.

A notable distinction of this study lies in its target population. Prior PS research has primarily focused on older adults or individuals with clinically diagnosed MCI, populations where cognitive decline is already measurable, and intervention effects are more readily detectable [19–21]. In contrast, the present trial enrolled a broader spectrum of healthy, working-age adults (35-65) who reported only subjective cognitive decline without objective impairment. It is plausible that cognitive effects in this healthier, higher-functioning population are more subtle and domain-specific than those observed in impaired cohort. This may explain why composite memory, a broad measure of verbal and visual recall, did not show a statistically significant change, whereas more specific domains such as working memory and motor speed showed possible domain-specific effects in exploratory analyses. This pattern also reflects a wider phenomenon in which an intervention may show limited efficacy once substantial impairment is present but offers clearer advantages when used preventively before measurable decline occurs. A well-known parallel exists in dementia research on omega-3 fatty acids. Meta-analyses of randomized controlled trials show that DHA/EPA supplementation has little therapeutic effect in individuals with established AD or moderate cognitive impairment [22], whereas longitudinal and epidemiological studies consistently associate higher baseline omega-3 status and long-term intake with a reduced risk of developing cognitive decline or dementia [23]. Notably, the inclusion of the (EPA + DHA)/AA fatty-acid ratio as a time-varying covariate in the exploratory mixed-effects models also raises the possibility that baseline nutritional status may modulate cognitive responsiveness. Participants with lower baseline omega-3–related profiles may have had greater capacity for improvement, consistent with the possibility that PS exerts its strongest effects in individuals who begin with suboptimal functional reserves. This prevention–treatment asymmetry suggests that PS may exert its most meaningful effects by supporting cognitive resilience in individuals with suboptimal baseline status, while in healthier high-functioning populations its benefits may be more localized to specific executive or attentional processes rather than broad cognitive restoration. Nonetheless, APOE ε4 genotype was not assessed in this study. Because APOE status may influence the relationship between omega-3 status and cognitive outcomes, residual confounding related to genetic susceptibility cannot be fully excluded.

The finding for working memory in the exploratory analysis (*p* = 0.037) is of particular interest for this study population. Unlike composite memory score, which reflects general recall ability, working memory represents a core executive function responsible for the active retention and manipulation of information in service of cognitive goals. This function is crucial for real-world demands of planning, problem-solving, and decision-making faced by healthy, working-age adults [24]. This exploratory finding suggests that BioPS^®^ may help support these demanding executive processes rather than improving general recall in a population that is not yet impaired. This domain-specific pattern in a healthy cohort is consistent with other nutritional interventions, such as *Bacopa monnieri*, which has also been shown to enhance complex cognitive processing speed rather than simple memory recall in healthy adults [25]. Furthermore, the exploratory finding for motor speed (*p* = 0.005) is noteworthy not only for the performance improvement itself, but also for its apparent moderation by the participant’s blood fatty acid ratio profile. This three-way interaction is consistent with the “sub-optimal status” hypothesis, suggesting a plausible biological mechanism. This exploratory interaction suggests that the observed motor speed response may have varied according to omega-3-related nutritional status, although this finding should be interpreted cautiously given the two-timepoint design and exploratory nature of the analysis. The result implies that the supplement’s effect on neuromotor processing speed may not be uniform across individuals but is influenced by underlying nutritional status producing the strongest benefits in those starting from lower omega-3–related baseline. This concept of nutrient-nutrient interaction is well-established; a strong parallel is seen in studies where B vitamins, which lower homocysteine, only succeeded in slowing brain atrophy in participants who also had a high baseline omega-3 status [26]. This precedent supports the possibility that the efficacy of a nutritional supplement can be highly dependent on the individual’s broader nutritional profile.

The exploratory mixed-effects analysis provided an additional way to examine possible domain-specific change patterns and the potential moderating role of nutritional status in this healthy cohort. Because the prespecified primary ANCOVA did not detect a significant effect on the primary endpoint, these exploratory observations should be interpreted cautiously. Nonetheless, they may help inform the design of future nutrition-cognition trials in which effect sizes are expected to be modest and inter-individual variability substantial. All in all, the study’s findings must be interpreted in the context of several limitations. First, the sample size was powered for the composite memory endpoint, and the study was likely underpowered to detect the more subtle, domain-specific effects observed in working memory and motor speed. Second, these findings were identified through exploratory mixed-effects modeling rather than the prespecified primary analysis; therefore, confirmation in future trials specifically designed and powered for these domains is needed. Third, normal cognition was screened using an MMSE threshold of ≥24 as specified in the original protocol, but education-adjusted MMSE thresholds were not applied. Although participants also completed the Cognitrax® battery at screening, some influence of education on cognitive classification cannot be fully excluded. Fourth, the study population consisted exclusively of healthy Japanese adults, which may limit generalizability to other ethnic groups or populations with different dietary patterns. Finally, the inclusion criterion of subjective memory decline is inherently heterogeneous, potentially introducing variability that may have diluted the ability to detect broader treatment effects.

## 5. Conclusion

In conclusion, daily supplementation with 300 mg of soybean-derived PS was safe and well tolerated in healthy, middle-aged adults with subjective memory complaints. The prespecified primary efficacy analysis did not show a significant benefit for composite memory at week 12. However, exploratory secondary analyses suggested the possibility of domain-specific effects in working memory and motor speed and indicated that nutritional status may influence cognitive responsiveness. These exploratory findings require confirmation in larger, prospectively designed studies.

## Data Availability

All data produced in the present work are contained in the manuscript

## CRediT authorship contribution statement

Yin Liu: Writing – original draft, Methodology, Formal analysis; Yunping Tang: Writing – review & editing, Conceptualization; Galex K.S. Neoh: Writing – original draft, Methodology; Yangfan Lu: Methodology, Formal analysis; Tsuyoshi Takara: Methodology, Formal analysis; Su Jiang: Writing – review & editing, Funding acquisition.

## Acknowledgement

The authors gratefully acknowledge all study participants and the staff members involved in conducting the study and collecting the study data.

## Data availability statement

The data that supports the findings of this study are not publicly available because they contain individual participant information and are subject to ethical and privacy restrictions under the original institutional review board approval. De-identified data may be made available from the corresponding author, Su Jiang, upon reasonable request, provided that the request is accompanied by appropriate institutional review or data-sharing agreements to ensure confidentiality and compliance with applicable ethical guidelines.

